# Cost-Effectiveness and Cost-Utility of a Colon Capsule Endoscopy in a Population-Based Screening Program for Colorectal Cancer

**DOI:** 10.64898/2026.05.30.26354522

**Authors:** Gerard Carot-Sans, Anastasios Koulaouzidis, Aitor Gonzalez-Amezcua, Ulrik Deding, Konstantinos Triantafyllou, Dan Ouchi, Bettina Eriksen, Benedicte Schelde-Olesen, Gunnar Baatrup, Jordi Piera-Jiménez, Claudia Erika Delgado Espinoza, Claus Duedal Pedersen, Angus J M Watson, Ferran Torres, Caridad Pontes

**Affiliations:** Catalan Health Service, Barcelona, Spain; Digitalization for the Sustainability of the Healthcare System (DS3), Barcelona, Spain; University of Southern Denmark, Odense, Denmark; Odense University Hospital, Svendborg, Denmark; Universitat Autònoma de Barcelona, Cerdanyola del Vallès, Spain; National and Kapodistrian University of Athens, Athens, Greece; Attikon University General Hospital, Athens, Greece; Stratos AI Aps, Odense, Denmark; openEHR International, Cardiff, United Kingdom; Fundació TICSalut i Social, Barcelona, Spain; Clinical Pharmacology Service, Hospital de la Santa Creu i Sant Pau, Barcelona, Spain; Institute of Applied Health Sciences, University of Aberdeen, Aberdeen, United Kingdom

**Keywords:** Colon capsule endoscopy, colonoscopy, colorectal cancer, cost effectiveness, cost utility

## Abstract

**Background:** Colon capsule endoscopy (CCE) has been proposed as a non-invasive alternative to colonoscopy for colorectal cancer (CRC) screening, offering greater patient comfort and potentially reducing healthcare burden. However, its cost-effectiveness in population-based screening remains uncertain.

**Methods:** This study used a state-transition (Markov) model to simulate lifetime outcomes of CRC screening in Denmark, Scotland, and Spain, comparing the standard pathway based on fecal immunochemical testing (FIT) followed by colonoscopy with an alternative pathway replacing colonoscopy with CCE after a positive FIT result. The model incorporated costs (2024 euros), quality-adjusted life-years (QALYs), and CRC cases avoided, applying a yearly discount rate of 3%. Deterministic sensitivity analyses explored uncertainty in capsule cost, adherence, and reinvestigation rates for non-advanced polyps.

**Results:** Across all settings, CCE resulted in higher costs but slightly increased effectiveness and utility (mean QALYs 28.7 vs. 28.8; CRC detected 0.032–0.034 vs. 0.035–0.037 per person). Incremental cost-effectiveness ratios (ICER) ranged from €43,538 in Spain to €136,930 in Denmark per additional CRC detected. Capsule cost was the main driver of ICER variation, whereas adherence rates had minimal effect on cost-effectiveness. Changes in the prevalence of non-advanced polyps had a modest impact, except when capsule prices were high.

**Conclusions:** Overall, replacing colonoscopy with CCE slightly increases detection and health gains at the expense of higher costs. Cost-effectiveness largely depends on capsule price and adherence. Artificial intelligence-assisted CCE interpretation may further improve diagnostic and economic performance, potentially supporting adoption in large-scale CRC screening programs.

## Introduction

Colon capsule endoscopy (CCE) has been proposed as a non-invasive alternative to colonoscopy, flexible sigmoidoscopy, and computed tomographic colonography for exploring the large bowel [1–3]. CCE can be performed in an outpatient setting without the need for sedation - anesthesia, or specialized professional involvement. Consequently, CCE not only minimizes the risk of adverse events during the procedure but can also reduce patient inconvenience and alleviates the burden on healthcare systems [4–6].

Clinical trials evaluating the accuracy of CCE for detecting colorectal neoplasia have demonstrated its high performance, with greater sensitivity and specificity compared to computed tomographic colonography and colonoscopy [3,6,7]. Although CCE has seen limited adoption in real-world settings, patient-reported outcome measures from randomized controlled trials (RCTs) indicate that CCE is well-accepted by patients and equally preferred compared to conventional techniques [8]. As CCE technology continues to evolve, the recent incorporation of artificial intelligence (AI) algorithms for automated image analysis may further influence its diagnostic performance, workflow efficiency, and clinical implementation [9–11].

Taken together, the evidence to date suggests that CCE has significant potential to enhance most domains of the quadruple aim of healthcare value in the management of CRC, offering benefits in patient experience, effectiveness, and potentially healthcare professional satisfaction. However, the trade-off between costs and benefits remains underexplored. This trade-off is particularly critical in population-level screening programs, where small differences in cost-effectiveness can lead to significant financial implications when scaled. In this analysis, we used updated data on performance, adherence, and the natural history of CRC progression to simulate the cost, utility, and effectiveness of incorporating CCE into CRC screening programs.

## Methods

### Model overview

We developed a state-transition (i.e., Markov) model to assess the cost-utility and cost-effectiveness of a CRC screening pathway based on CCE compared with the current screening pathway which uses colonoscopy. The two pathways were developed using the TreeAge Pro software for a simulation analysis [12].

Figure 1 shows an overview of the two screening pathways; the entire model developed in TreeAge Pro is provided in Figure S1 as Supplementary material. First, we designed a reference screening pathway according to the recommendations of relevant guidelines on CRC screening in effect in Europe [13–16]. Briefly, for the reference pathway, we assumed that everyone aged 50 or older were invited to undergo a fecal immunochemical test (FIT) for the detection of blood in the stool every 2 years. Individuals testing negative in the first-round of FIT were invited to re-enter the screening pathway normally at 2 years, whereas those with positive FIT but no findings in subsequent colonoscopy or CCE were re-invited to enter the pathway after 10 years. Individuals with low-risk findings detected in colonoscopy or CCE (defined as polyps smaller than 6 mm or ≤3 adenomas smaller than 1 cm) were not actively surveilled but rather re-entered the routine screening program with biennial invitations. In contrast, individuals with advanced findings (One polyp larger than 6 mm or >3 adenomas smaller than 1 cm), underwent active surveillance with annual FIT test for 5 years, after which they returned to the general screening program, every 2 years.

**Figure 1.**
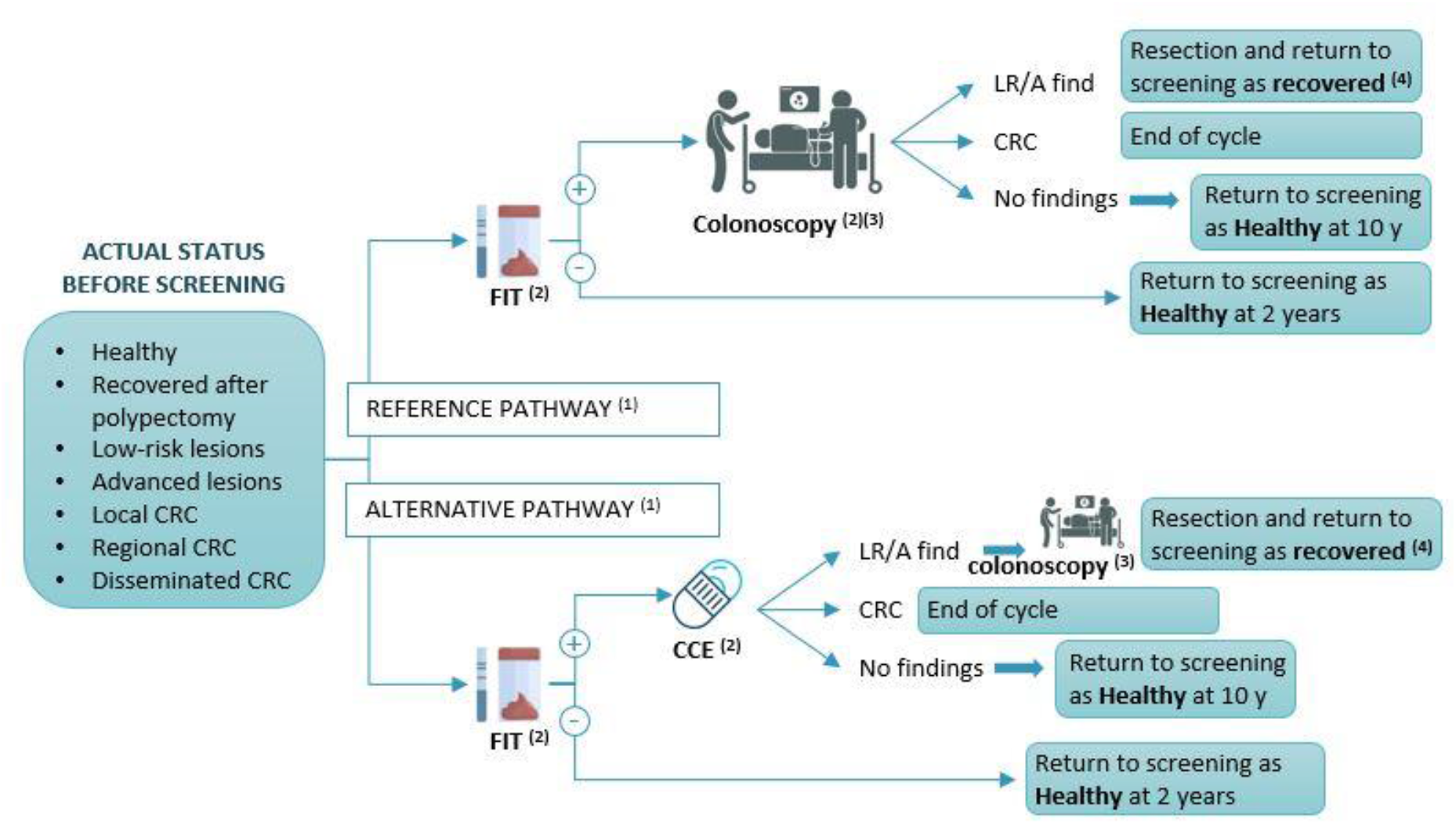
Overview of the two pathways for a systematic screening of colorectal cancer. **CCE**: colon capsule endoscopy. **CRC**: colorectal cancer **FIT**: fecal immunochemistry test. **LR/A find:** low-risk / advanced findings. **OC**: optical colonoscopy. ^(1)^Background mortality was accounted for at screening entry. ^(2)^When available, adherence to each test was considered. ^(3)^Adverse events were considered for optical colonoscopy. ^(4)^Follow-up interval times were based on findings: individuals with low-risk findings were re-invited to the program every 2 years; individuals with APL are actively surveilled with FIT yearly for 5 years and re-invited to the regular program every 2 years thereafter.

In the alternative pathway, we replaced colonoscopy with CCE following a positive FIT result. Based on guideline recommendations for removing polyps, irrespective of their risk, we introduced a colonoscopy following CCE in individuals with any polyp identified during CCE. In both pathways (i.e., reference and alternative), individuals exited the simulation upon CRC diagnosis, death, or after 50 yearly cycles. Background mortality was accounted for at the beginning of each cycle.

### Model inputs

State and transition probabilities were established based on values reported in the literature. Probabilities (provided in the Supplementary appendix along with the data sources) were grouped into five categories: background mortality, which accounted for overall mortality risk, irrespective of CRC (Table S1), probabilities associated with the epidemiology (starting prevalence) and natural history (risk of progression) of CRC (Table S2), performance of FIT, colonoscopy, and CCE for the different stages (Table S3), complications associated with colonoscopy (Table S4), and adherence to the different tests of the screening program (Table S5). All state and transition probabilities were assumed to be identical across the three countries, except for FIT adherence, for which country-specific data from Denmark were available [17] and therefore used (Table S5).

Costs were estimated separately on a country basis, and included program roll-out, visits, tests (FIT, CCE, screening colonoscopy, and colonoscopy with biopsy, polypectomies), complications and CRC treatment according to stage (Table S6). Given the potential variability of costs across Europe, we replicated the analysis by considering the values from three different countries with consolidated screening programs for CRC based on systematic FIT followed by colonoscopy: Denmark [18], Scotland [19], and Spain [20]. Costs were represented in 2024 euros.

### Endpoints and base-case analysis

For the cost-utility analysis, we computed the number of QALYs resulting from each pathway and estimated the incremental cost-utility rate. QALYs were retrieved from Ness et al. [21] and stratified according to CRC stage at diagnosis. The same utilities were considered for all countries.

Additionally, we conducted a cost-effectiveness analysis using the “rate of remaining CRC” endpoint proposed by Ludicame et al. [22] and defined as screened-and-undiagnosed plus unscreened-and-undiagnosed CRC. This was estimated based on the proportion between the number of CRC diagnosed and the incident CRC in each pathway. The incremental cost-effectiveness ratio (ICER) of this outcome represents the cost per additional CRC detected over the time-frame of the simulation (i.e., up to 50 years). This outcome was complemented with the breakdown description of the CRC detected and missed in the two pathways.

Other descriptive variables included the overall testing burden (i.e., cumulative number of tests or exams conducted on a patient), the per-diagnosis cost (i.e., the total cost divided by the number of CRC and advanced polyps detected), the number of polyps detected, the estimated missed CRC lethality (i.e., mortality among individuals with missed CRC), and the estimated CRC onset (i.e., the number of CRC appearing in each pathway according to the model).

The utility, effectiveness, and cost parameters were discounted by a yearly rate of 3%

### Sensitivity analyses

To evaluate the impact of uncertainty on the key parameters of the model, a deterministic sensitivity analysis was conducted on variables with high level of uncertainty, high heterogeneity in the literature, and/or expected variation as technological development advances. The analysis was implemented on the same microsimulations of the stochastic Markov model used in the base case scenario.

First, the price of the CCE was assumed to fluctuate between 50% lower values, driven by economies of scale, and 50% higher values, influenced by technological advancements. Second, owing to the early-stage implementation of the CCE, we found no reliable information regarding adherence rates to CCE exam in a real-world screening setting. Based on the non-invasiveness of the exam, we assumed a high adherence level for the base case analysis; the effect of lower (up to 70%) adherence to CCE after a positive finding in FIT was explored. Third, we found heterogeneous values for adherence to first and subsequent FIT assessments; therefore, the effect of variations in this parameter (30% to 80%) were explored. The probability of polypectomy due to low-risk findings was included in the sensitivity analysis to account for reinvestigation rates in screening programs that do not systematically remove diminutive or isolated polyps [23,24], as well as to reflect screening scenarios involving populations with different baseline risk levels (50% above and below the 24% used for the base case scenario). Since price variations were expected to be the most remarkable and impactful parameter, all other parameters were investigated along with price variations.

## Results

### Base-case analysis

The pathway based on the CCE was more costly than the reference pathway in all European countries, particularly Denmark, where the price of the capsule by far exceeded that of colonoscopy. The CCE pathway was associated with similar (slightly higher) QALY gains to the colonoscopy pathway (Table 1).

**Table 1.** Results for the base-case analysis.

|  | Cost (€) | Incremental<br>cost (€) | QALYs | Incremental<br>QALY | ICUR<br>(€/QALY) | Incident<br>CRC | Incremental<br>detection of<br>CRC | ICER<br>(€/CRC detected) |
| --- | --- | --- | --- | --- | --- | --- | --- | --- |
| **Denmark** |  |  |  |  |  |  |  |  |
| Colonosc.<br>Pathway | 946.11 |  | 28.7 |  |  | 0.034 |  |  |
| CCE Pathway | 1,444.53 | 498.43 | 28.8 | 0.1 | **4,079.08** | 0.037 | 0.0036 | **136,930.02** |
| **Scotland** |  |  |  |  |  |  |  |  |
| Colonosc.<br>Pathway | 548.86 |  | 28.7 |  |  | 0.032 |  |  |
| CCE Pathway | 799.80 | 250.94 | 28.8 | 0.1 | **1,961.51** | 0.035 | 0.0030 | **83,368.19** |
| **Spain** |  |  |  |  |  |  |  |  |
| Colonosc.<br>Pathway | 383.78 |  | 28.6 |  |  | 0.032 |  |  |
| CCE Pathway | 512.21 | 128.44 | 28.8 | 0.3 | **467.99** | 0.035 | 0.0030 | **43,537.82** |

CCE: colon capsule endoscopy. CRC: colorectal cancer. ICER: incremental cost-effectiveness ratio. ICUR: incremental cost-utility rate. QALY: quality-adjusted life years.

The number of CRC detected were slightly higher in the CCE pathway (3.5 to 3.7 per 100 individuals, depending on the country) than the colonoscopy (3.2 to 3.4 per 100 individuals) (average relative difference 8%). Accordingly, in the colonoscopy pathway more CRC were missed, leading to higher values of incident CRC. The resulting ICER, presented as € per additional CRC detected over 50 years in the CCE compared with the colonoscopy pathway, ranged from 43,537.8 in Spain to 136,930.02 in Denmark (Table 1).

Table 2 summarizes other estimates of interest regarding healthcare resource utilization and health impact of the two pathways. In line with the overall cost estimate, each diagnosis was more expensive in the CCE pathway. However, the colonoscopy-based strategy entailed a substantially higher number of colonoscopies, as these included both exploratory and therapeutic (biopsy/excision) procedures, whereas in the CCE-based pathway, colonoscopy was largely restricted to confirmatory and therapeutic purposes following capsule findings. In contrast, FIT use and the number of therapeutic colonoscopies were broadly similar between approaches, indicating comparable downstream intervention needs once lesions were identified. In terms of diagnostic performance, the CCE pathway consistently detected more polyps per patient than the colonoscopy-based strategy. This enhanced detection was associated with earlier identification of CRC and translated into improved clinical outcomes, including fewer CRC-related deaths and a reduction in the number of undiagnosed CRC cases per 1,000 individuals. Overall, despite shifting the distribution of procedures, the CCE-based pathway achieved a higher diagnostic yield and improved effectiveness in reducing missed cancers and associated mortality.

**Table 2.** Additional health and resource use outcomes.

|  | **CCE**<br>**pathway** | **Colonoscopy**<br>**pathway** | Absolute<br>difference<sup>(1)</sup> | Relative<br>difference<sup>(2)</sup> |
| --- | --- | --- | --- | --- |
| **Test burden** (*tests and/or exams per individual*) |  |  |  |  |
| Overall test burden |  |  |  |  |
| Denmark | 8.35 | 7.35 | 1.01 | 14% |
| Scotland and Spain | 8.79 | 7.73 | 1.06 | 14% |
| FIT |  |  |  |  |
| Denmark | 7.69 | 7.00 | 0.69 | 10% |
| Scotland and Spain | 8.11 | 7.37 | 0.74 | 10% |
| Exploratory colonoscopies<sup>(3)</sup> |  |  |  |  |
| Denmark |  | 0.16 |  |  |
| Scotland and Spain |  | 0.17 |  |  |
| Colonoscopy with biopsy/excision |  |  |  |  |
| Denmark | 0.22 | 0.18 | 0.04 | 23% |
| Scotland and Spain | 0.23 | 0.19 | 0.04 | 22% |
| Exploratory CCE |  |  |  |  |
| Denmark | 0.44 |  |  |  |
| Scotland and Spain | 0.45 |  |  |  |
| **Individual cost** |  |  |  |  |
| Per diagnosis cost (*€/patient*) |  |  |  |  |
| Denmark | 515.03 | 454.44 | 60.59 | 13% |
| Scotland | 433.17 | 383.06 | 50.11 | 13% |
| Spain | 336.77 | 293.53 | 43.24 | 15% |
| **Diagnostic performance** |  |  |  |  |
| Polyps detected (*No./patient*) |  |  |  |  |
| Denmark | 0.19 | 0.15 | 0.04 | 26% |
| Scotland and Spain | 0.20 | 0.16 | 0.04 | 24% |
| CRC onset (*average per patient*) |  |  |  |  |
| Denmark | 0.054 | 0.062 | -0.0085 | -14% |
| Scotland and Spain | 0.052 | 0.059 | -0.0072 | -12% |
| CRC deaths (*average per 1,000 patients*) |  |  |  |  |
| Denmark | 23.4 | 29.61 | -6.21 | -21% |
| Scotland and Spain | 20.9 | 27.76 | -6.86 | -25% |
| Undiagnosed CRC (*average per 1,000 individuals*) |  |  |  |  |
| Denmark | 178.21 | 229.94 | -51.73 | -22% |
| Scotland and Spain | 160.37 | 209.76 | -49.39 | -24% |

State and transition probabilities were the same for Scotland and Spain; specific values for FIT adherence were applied to Denmark and, therefore, results are presented separately.
**CCE:** colon capsule endoscopy. **CRC:** colorectal cancer.
<sup>(1)</sup> Per 1,000 patients.
<sup>(2)</sup> Using colonoscopy as a reference.
<sup>(3)</sup> Colonoscopies that did not result in excision or biopsy and where, therefore, optical exploration only.

The descriptive analysis of the contribution of each factor (e.g., FIT adherence and performance, CCE/colonoscopy adherence and performance) to undetected cancers throughout simulation cycles revealed that low adherence to colonoscopy and false negative exams were the factors with highest contribution to omitted CRC (Figure S2).

### Sensitivity analysis

Figure 2 summarizes the results of the two-way deterministic sensitivity analyses exploring the impact of CCE unit cost in combination with key model parameters (i.e., FIT adherence, CCE adherence, and prevalence of low-risk lesions) on ICER in the three countries. Variations in the key model parameters had higher impact in Denmark, where the cost of the CCE was highest. Also, across all panels, increasing CCE cost was consistently associated with higher ICER values in all three countries. This gradient was observed irrespective of the second parameter varied (FIT adherence, CCE adherence, or prevalence of low-risk findings), indicating that CCE unit price was a dominant driver of ICER outcomes. For a given cost of the CCE, increasing values of adherence to FIT were associated with higher ICER, with a maximum at 80% adherence. Variations in adherence to CCE had a modest effect on ICER, except for very low values (i.e., 70%), which resulted in a sharp increase in ICER across all CCE costs. Finally, variations in the prevalence of non-advanced polyps showed alternate patterns with relatively small variations in ICER, except for the highest CCE costs.

**Figure 2.**
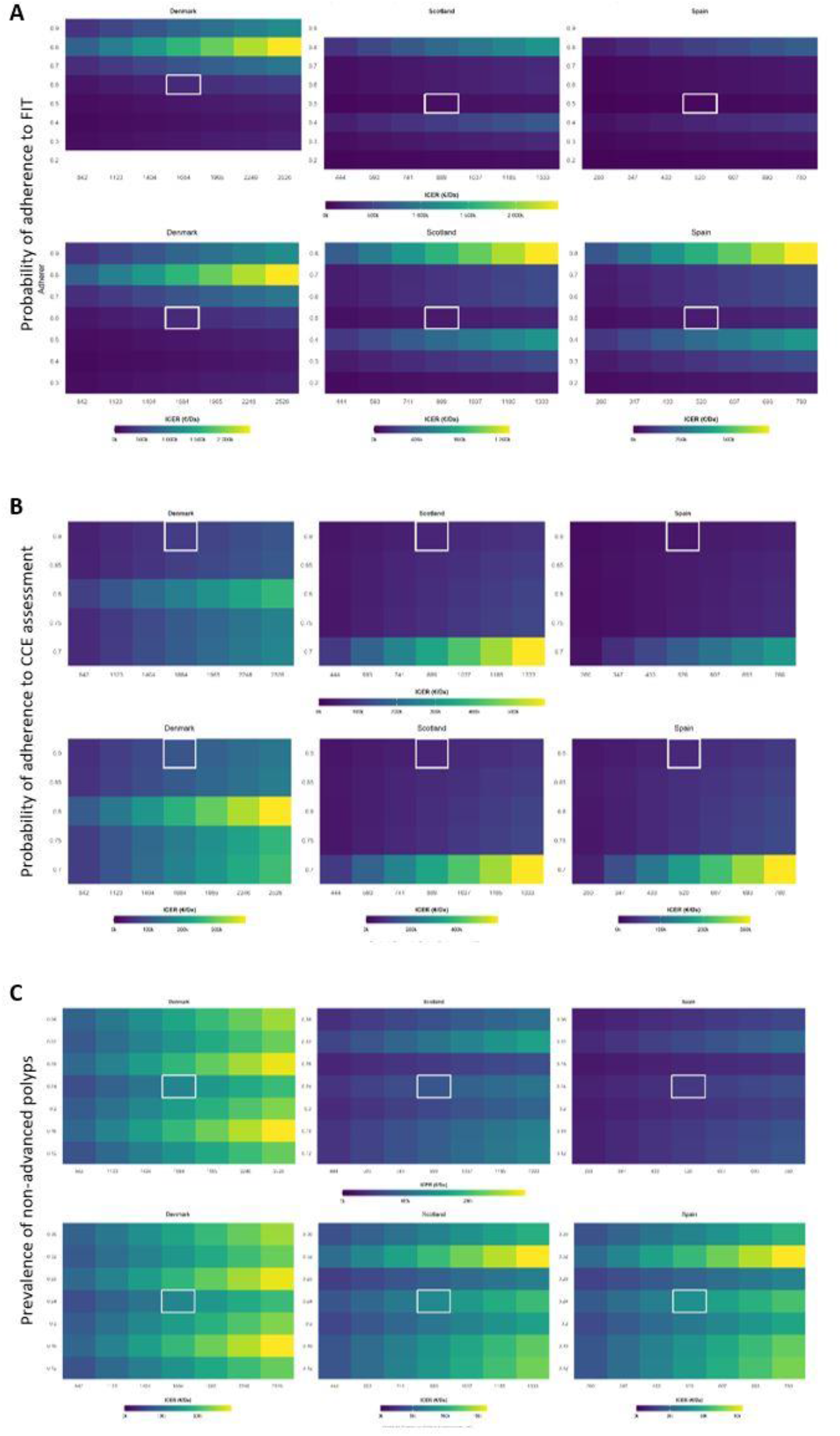
Two-way deterministic sensitivity analysis for the incremental cost-effectiveness ratio (ICER, expressed in € per colorectal cancer detected) according to variation in parameters with low uncertainty or high risk of fluctuation: **A:** adherence to fecal immunochemistry test (FIT) and cost of CCE. **B:** adherence to CCE after a positive FIT result and cost of CCE. **C:** prevalence of non-advanced polyps and cost of CCE. For each set of multicounty analyses, plots are represented using two scales: same scale for all countries (panel lines above) and scale optimal scale for each country (panel lines below).

Similarly to ICER results, ICUR was primarily affected by changes in the cost of CCE. The effect changes in the probability of adherence to FIT and CCE was limited, except for Denmark, where low values of adherence to CCE were translated to higher ICUR values (Figure 3). Variations in the prevalence of non-advanced polyps resulted in alternate patterns of increase and decrease of ICUR.

**Figure 3.**
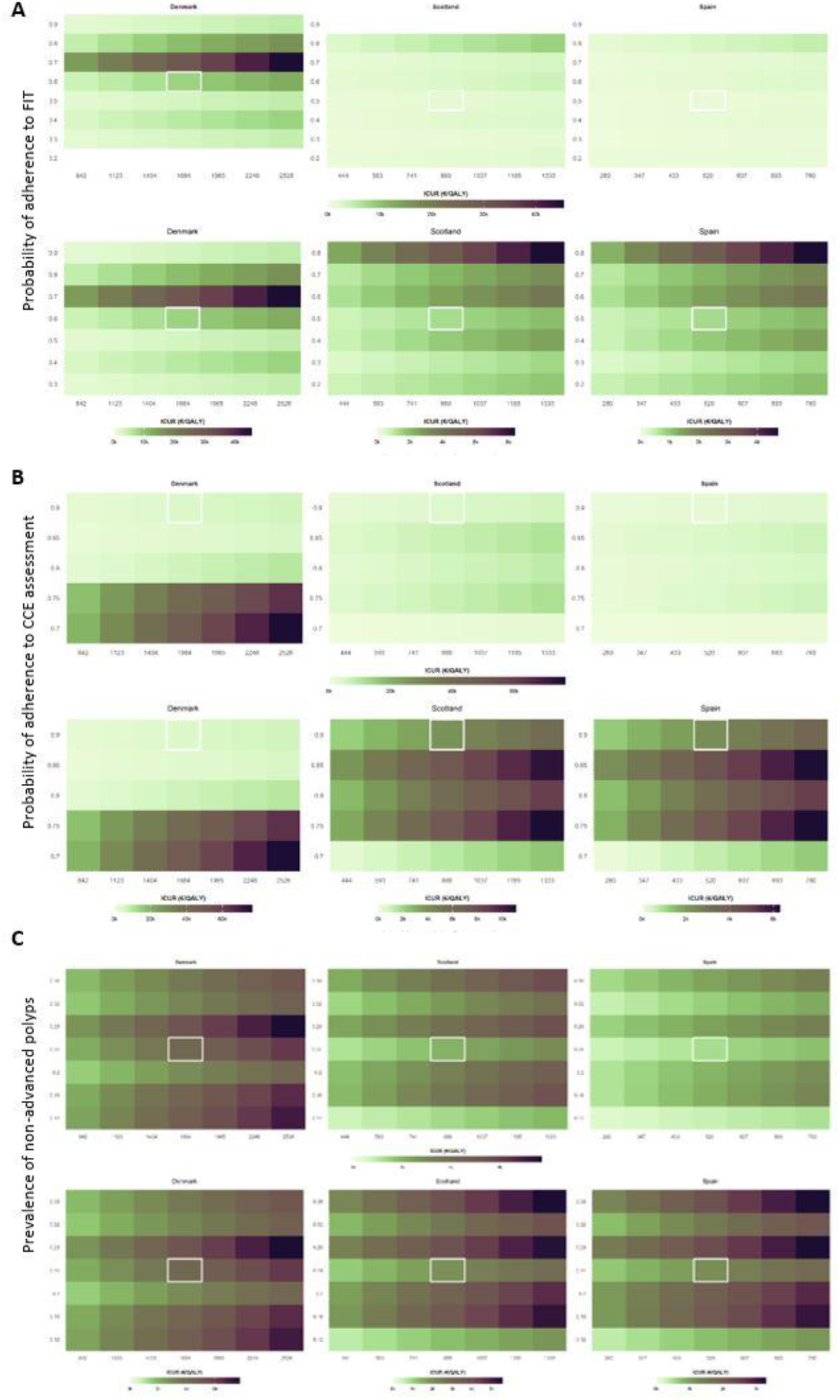
Two-way deterministic sensitivity analysis for the incremental cost-utility ratio (ICUR, expressed in € per quality-adjusted life years gained) according to variation in parameters with low uncertainty or high risk of fluctuation: **A:** adherence to fecal immunochemistry test (FIT) and cost of CCE. **B:** adherence to CCE after a positive FIT result and cost of CCE. **C:** prevalence of non-advanced polyps and cost of CCE. For each set of multicounty analyses, plots are represented using two scales: same scale for all countries (panel lines above) and scale optimal scale for each country (panel lines below).

## Discussion

In this simulation of CRC screening pathways at the population level, we found that replacing colonoscopy with CCE resulted in higher costs and moderately higher utility (QALYs) and effectiveness (CRC early detected and potentially avoided) than the conventional strategy based on colonoscopy. The sensitivity analysis with variations in adherence to CCE and the price of the capsule revealed that increased prices of the capsule had a stronger impact on ICER and ICUR. Extreme values of adherence to FIT screening and CCE assessment influenced the ICER, which remained relatively unchanged for moderate changes in this parameter, particularly in Scotland and Spain, were costs of CCE were remarkably low. Changes in the prevalence of non-advanced polyps resulted in alternating patterns of ICER and ICUR, reflecting the high complexity of the trade-off between higher number of repeated exams (i.e., colonoscopy after low-risk findings in CCE) and the effectiveness.

The high sensitivity of CCE for detecting polyps has been highlighted in various studies, underscoring its potential as an effective tool for CRC screening [25–27]. Among the most comprehensive evaluations of this strategy is the ScotCap registry, which reported a per-polyp sensitivity of 97.1% for detecting polyps measuring 6 mm or larger and 95.8% for those 10 mm or larger [25,26]. However, the authors raised a critical concern regarding the potential for CCE to “over-report” polyps, as some polyps identified during CCE were not subsequently confirmed during follow-up colonoscopy. This discrepancy, which is particularly common in smaller polyps [28,29], may be attributed to factors such as false positives, double reporting, differences in bowel preparation quality, or challenges in correlating capsule findings with precise anatomical locations during colonoscopy. While the probability of false-positive findings in CCE has not yet been thoroughly investigated (and, therefore, could not be incorporated into our simulation), specificity will be a key factor in ensuring the cost-effectiveness of the capsule.

One of the challenging aspects of integrating CCE into screening or diagnostic pathways for colorectal cancer is the need for re-investigation with colonoscopy after a positive finding in a CCE exam. In our simulation, this scenario was modeled by performing a colonoscopy whenever any finding was detected during the CCE, regardless of the associated risk. This approach aligns with current guidelines, which consistently recommend the removal or biopsy of polyps of any risk detected during a colonoscopy examination [13–15]. However, diminutive polyps rarely progress to cancer [30], and there is growing recognition in the field that such polyps may not always require removal; instead, advancements in imaging techniques capable of distinguishing between adenomas and non-adenomatous polyps have been proposed as a potential alternative to routine excision or biopsy [31]. Alternatively, some screening programs do not mandate systematic removal of diminutive polyps [24]. To account for variability in re-investigation criteria, which are expected to evolve as *in situ* polyp characterization through the use of AI enables more accurate risk stratification, we conducted a sensitivity analysis using different prevalence values for non-advanced polyps (serving as a proxy for re-investigation with colonoscopy in our model). The results showed that the ICER was only marginally affected at low CCE costs, suggesting that, although re-investigation is relevant, cost-effectiveness remains primarily driven by the cost of the CCE procedure. Of note, an alternate pattern of ICER values across prevalences of non-advanced polyps at a given CCE cost suggests high complexity of the trade-off between the number of early CRC detected and potentially avoided (which reduces the ICER) and the increased number of examinations after CCE (which increases the ICER).

Re-investigation with colonoscopy in real-world practice has implications beyond cost-effectiveness, including increased workload, delays in diagnosis and intervention compared with initial colonoscopy, and the need for repeated bowel preparation, which is burdensome for patients. Therefore, despite the overall favorable cost-effectiveness profile of the CCE pathway, there is a clear need to develop screening strategies that minimize the likelihood of re-investigation among individuals selected for CCE before considering this technique in the screening setting [32].

In our simulation, the re-examination process following a positive finding in CCE contributed to a higher test burden compared to the colonoscopy pathway; an increased number of FIT tests further added to this burden. Consequently, the larger volume of tests led to higher costs per diagnosed patient. On the other hand, the CCE pathway demonstrated an advantage by identifying more polyps at earlier stages, effectively preventing their progression to CRC. Overall, considering the transition probabilities incorporated into the model for the natural history of CRC, this improved detection rate resulted in lower lethality for the CCE pathway compared to the colonoscopy pathway. According to the sensitivity analysis, this trend was not severely affected when considering a range of adherence rates to CCE examination.

With recent advancements in artificial intelligence, the integration of deep learning algorithms into CCE has been proposed as a transformative enhancement [10]. These algorithms are capable of detecting lesions in the large bowel with remarkable accuracy. By leveraging AI, the workload of endoscopists could be significantly reduced, as they would only need to review AI-selected images rather than the entire image sequence [10]. Additionally, this technology could minimize inter-operator variability, ensuring more consistent results. Moreover, AI-integrated systems can deliver results in near real-time. This capability opens the door to designing efficient care pathways, where patients with positive findings from a CCE exam could proceed directly to colonoscopy without the need for additional bowel preparation. Although this strategy is still under development, it holds the potential to revolutionize CRC screening by enhancing efficiency and, therefore, cost-effectiveness, as well as by increasing patient comfort and acceptability.

Since AI integration is still in a developmental phase, we were unable to include a third pathway for evaluating the cost-effectiveness of AI-enhanced CCE. This approach is expected to be highly expensive during the initial stages of implementation due to the need to recover development investments. However, costs are likely to decrease significantly as adoption scales up. Additionally, the reduction in reading time required from endoscopists is expected to enhance the cost-effectiveness balance over time. Besides the inability to evaluate the cost-effectiveness of AI-enhanced CCE, our results were also limited by certain heterogeneity in performance metrics, which varied across source studies, particularly due to differences in polyp size. Moreover, we did not account in the model for the potential loss of effectiveness associated with inadequate bowel preparation, a critical factor for the optimal functioning of CCE. Despite these limitations, our model was significantly more comprehensive than most published studies evaluating the cost-effectiveness of CRC screening strategies, that tend to oversimplify the complexity of decision-making and the sequential tests encountered in routine public health practice.

In summary, CCE holds substantial promise for CRC detection. Nevertheless, two key factors may limit its widespread adoption in CRC screening. First, the current costs associated with CCE must be reduced for this approach to truly transform screening practice. Second, although re-investigation had a limited impact on ICER at lower CCE costs, the drawbacks of frequent re-investigations highlight the need to develop screening pathways that identify candidates with a low likelihood of requiring follow-up colonoscopy before the systematic implementation of CCE. The integration of AI into CCE, though still in its developmental phase, has the potential to be a game changer, significantly enhancing both the utility and cost-effectiveness of this technology.

## Data Availability

All data utilized and generated in this study are aggregate data derived from publicly available repositories and published literature, as detailed in the manuscript and its supplementary material. No individual-level patient data were used.

## Abbreviations

AI: Artificial intelligence.
CCE: Colon capsule endoscopy.
CRC: colorectal cancer.
FIT: fecal immunochemical test
ICER: incremental cost-effectiveness ratio
QALY: Quality-adjusted life years
RCT: Randomized-controlled trial

## Acknowledgements

The authors kindly thank Andrew Munzer (TreeAge Pro) for his insightful comments on study results, and Louise McCullagh, Leah Bohn, and Tine Mikkelsen for the management and coordination of the AICE project. Although the study was funded by the European Union, views and opinions expressed are those of the author(s) only and do not necessarily reflect those of the European Union or the European Commission. Neither the European Union or the European Commission can be held responsible for them.

## Author contributions

The study was initially conceived by GC-S, AK, FT, and CP. Literature review and selection of model inputs were conducted by AK, KT, FT, and CP. Analysis was conducted by AG-A and FT. GC-S, AG-A, FT, and CP wrote the first draft of the manuscript, which was further revised and edited by GC-S, AK, AG-A, UD, KT, DO, BE, BS-O, GB, JP-J, CEDE, CDP, AJW, FT, and CP. The final version was approved by all co-authors, who agreed to submit the manuscript for publication.

## Declarations

### Ethics Approval and Consent to Participate

Not applicable. This study is based on a mathematical state-transition simulation model utilizing aggregated, previously published secondary data, and did not involve any direct interaction with human participants or the collection of identifiable personal health information. Consequently, institutional review board approval and informed patient consent were not required.

### Funding Sources

This study was conducted as part of the project AICE, funded by the European Union under the HORIZON program (Grant Agreement No. 101057400) and co-funded by the United Kingdom Government.

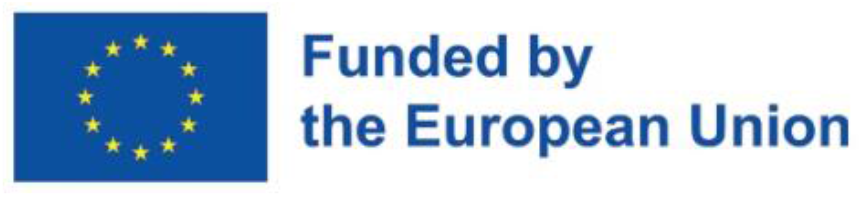

### Declaration of Competing Interests

Anastasios Koulaouzidis is a co-founder/shareholder of AJM Med-i-caps Ltd and has also been a co-founder and director of iCERV Ltd. He is a consultant to Jinshan Science & Technology Ltd. He has received research support (grants) from Given Imaging Ltd under the auspices of the European Society of Gastrointestinal Endoscopy (ESGE), and from IntroMedic (also referred to as SynMed/IntroMedic). He has received consultancy fees from Jinshan and lecture honoraria from Covidien/Medtronic. Additionally, he has benefited from lecture honoraria and educational travel support from a range of entities including Jinshan, Dr Falk Pharma UK, Ferring, Aquilant, and Almirall, and has participated in advisory board activities for companies such as Tillots, Ankon, and Dr Falk Pharma UK.

Bettina Eriksen and Claus Duedal Pedersen are members of Stratos AI, a spin-out company of the University of Southern Denmark and Odense University Hospital developing technologies for AI-powered screening and diagnosis, and which is partly owned by Science Ventures, representing SDU and Region Syddanmark (Rsyd).

All other authors declare that they received funding from their institutions and have no known competing financial interests or personal relationships that could have appeared to influence the work reported in this paper.

## References

1. Thygesen MK, Baatrup G, Petersen C, Qvist N, Kroijer R, Kobaek-Larsen M. Screening individuals’ experiences of colonoscopy and colon capsule endoscopy; a mixed methods study. Acta Oncol (Madr). 2019;58:S71–6. 10.1080/0284186X.2019.1581372

2. Chetcuti Zammit S, Sidhu R. Capsule endoscopy – Recent developments and future directions. Expert Rev Gastroenterol Hepatol. 2021;15:127–37. 10.1080/17474124.2021.1840351

3. González-Suárez B, Pagés M, Araujo IK, Romero C, Rodríguez De Miguel C, Ayuso JR, et al. Colon capsule endoscopy versus CT colonography in FIT-positive colorectal cancer screening subjects: A prospective randomised trial -The VICOCA study. BMC Med. BioMed Central Ltd; 2020;18. 10.1186/s12916-020-01717-4

4. Deding U, Bøggild H, Kaalby L, Hjelmborg J, Kobaek-Larsen M, Thygesen MK, et al. Socioeconomic differences in discrepancies between expected and experienced discomfort from colonoscopy and colon capsule endoscopy. Heliyon. 2024;10:e34274. 10.1016/j.heliyon.2024.e34274

5. Deding U, Bøggild H, Baatrup G, Kaalby L, Hjelmborg J, Thygesen MK, et al. Socioeconomic differences in expected discomfort from colonoscopy and colon capsule endoscopy. Prev Med (Baltim). 2023;173:107593. 10.1016/j.ypmed.2023.107593

6. Kjølhede T, ølholm AM, Kaalby L, Kidholm K, Qvist N, Baatrup G. Diagnostic accuracy of capsule endoscopy compared with colonoscopy for polyp detection: systematic review and meta-analyses. Endoscopy. 2021;53:713–21. 10.1055/a-1249-3938

7. Cash BD, Fleisher MR, Fern S, Rajan E, Haithcock R, Kastenberg DM, et al. Multicentre, prospective, randomised study comparing the diagnostic yield of colon capsule endoscopy versus CT colonography in a screening population (the TOPAZ study). Gut. 2021;70:2115–22. 10.1136/gutjnl-2020-322578

8. Deding U, Valdivia PC, Koulaouzidis A, Baatrup G, Toth E, Spada C, et al. Patient-reported outcomes and preferences for colon capsule endoscopy and colonoscopy: A systematic review with meta-analysis. Diagnostics. Multidisciplinary Digital Publishing Institute (MDPI); 2021. 10.3390/diagnostics11091730

9. Halvorsen N, Hassan C, Correale L, Pilonis N, Helsingen LM, Spadaccini M, et al. Benefits, burden, and harms of computer aided polyp detection with artificial intelligence in colorectal cancer screening: microsimulation modelling study. BMJ Medicine. BMJ Publishing Group; 2025;4. 10.1136/bmjmed-2025-001446

10. Nadimi ES, Braun JM, Schelde-Olesen B, Khare S, Gogineni VC, Blanes-Vidal V, et al. Towards full integration of explainable artificial intelligence in colon capsule endoscopy’s pathway. Sci Rep. 2025;15:5960. 10.1038/s41598-025-89648-z

11. Schelde-Olesen B, Herp J, Braun J-M, Koulaouzidis A, Bjørsum-Meyer T, Kaalby L, et al. Interobserver agreement between an artificial intelligence algorithm and colon capsule endoscopy readers on bowel-cleansing quality. iGIE. 2023;2:148–153.e3. 10.1016/j.igie.2023.04.006

12. TreeAge Software LLC. TreeAge Pro [Internet]. 2025 [cited 2026 May 11]. https://www.treeage.com/. Accessed 11 May 2026

13. Argilés G, Tabernero J, Labianca R, Hochhauser D, Salazar R, Iveson T, et al. Localised colon cancer: ESMO Clinical Practice Guidelines for diagnosis, treatment and follow-up†. Annals of Oncology. Elsevier Ltd; 2020;31:1291–305. 10.1016/j.annonc.2020.06.022

14. Lieberman DA, Rex DK, Winawer SJ, Giardiello FM, Johnson DA, Levin TR. Guidelines for colonoscopy surveillance after screening and polypectomy: A consensus update by the us multi-society task force on colorectal cancer. Gastroenterology. W.B. Saunders; 2012;143:844–57. 10.1053/j.gastro.2012.06.001

15. Von Karsa L, Patnick J, Segnan N, Atkin W, Halloran S, Lansdorp-Vogelaar I, et al. European guidelines for quality assurance in colorectal cancer screening and diagnosis: Overview and introduction to the full Supplement publication. Endoscopy. 2013. p. 51–9. 10.1055/s-0032-1325997

16. Hassan C, Antonelli G, Dumonceau J-M, Regula J, Bretthauer M, Chaussade S, et al. Post-polypectomy colonoscopy surveillance: European Society of Gastrointestinal Endoscopy (ESGE) Guideline – Update 2020. Endoscopy. 2020;52:687–700. 10.1055/a-1185-3109

17. National Healthcare Quality Institute (Denmark). Danish Colorectal Cancer Screening Database (DTS) [Internet]. Annual report. 2024 [cited 2026 Apr 23]. https://www.sundk.dk/media/auioalde/dts-aarsrapport_2024_offentliggjort_version_20251212.pdf. Accessed 23 Apr 2026

18. Durhuus JA, Galanakis M, Maltesen T, Therkildsen C, Rosthøj S, Klarskov LL, et al. A registry-based study on universal screening for defective mismatch repair in colorectal cancer in Denmark highlights disparities in screening uptake and counselling referrals. Transl Oncol. Neoplasia Press, Inc.; 2024;46. 10.1016/j.tranon.2024.102013

19. Mansouri D, McMillan DC, Crearie C, Morrison DS, Crighton EM, Horgan PG. Temporal trends in mode, site and stage of presentation with the introduction of colorectal cancer screening:A decade of experience from the West of Scotland. Br J Cancer. Nature Publishing Group; 2015;113:556–61. 10.1038/bjc.2015.230

20. Garcia M,Maria Borràs J, Binefa G, Milà N, Alfons Espinàs J, Moreno V. Repeated screening for colorectal cancer with fecal occult blood test in Catalonia, Spain. European Journal of Cancer Prevention. 2012;21:42–5. 10.1097/CEJ.0b013e32834a7e9b

21. Ness RM, Holmes AM, Klein R, Dittus R. Utility valuations for outcome states of colorectal cancer. Am J Gastroenterol [Internet]. 1999;94:1650–7. 10.1111/j.1572-0241.1999.01157.x

22. Lucidarme O, Cadi M, Berger G, Taieb J, Poynard T, Grenier P, et al. Cost-effectiveness modeling of colorectal cancer: Computed tomography colonography vs colonoscopy or fecal occult blood tests. Eur J Radiol. 2012;81:1413–9. 10.1016/j.ejrad.2011.03.027

23. Kaalby L, Deding U, Kobaek-Larsen M, Havshoi A-LV, Zimmermann-Nielsen E, Thygesen MK, et al. Colon capsule endoscopy in colorectal cancer screening: a randomised controlled trial. BMJ Open Gastroenterol. 2020;7:e000411. 10.1136/bmjgast-2020-000411

24. Deding U, Bjørsum-Meyer T, Kaalby L, Kobaek-Larsen M, Thygesen MK, Madsen JB, et al. Colon capsule endoscopy in colorectal cancer screening: Interim analyses of randomized controlled trial CareForColon2015. Endosc Int Open. 2021;09:E1712–9. 10.1055/a-1546-8727

25. MacLeod C, Hudson J, Brogan M, Cotton S, Treweek S, MacLennan G, et al. ScotCap – A large observational cohort study. Colorectal Disease. John Wiley and Sons Inc; 2022;24:411–21. 10.1111/codi.16029

26. MacLeod C, Rajapaksha N, Brown C, Hudson J, Asif Z, Watson AJM, et al. The ScotCap registry: An evaluation of 1000 colon capsule endoscopy procedures carried out in Scotland. Colorectal Disease. John Wiley and Sons Inc; 2025;27. 10.1111/codi.17271

27. MacLeod C, Rajapaksha N, Brown C, Hudson J, Asif Z, Watson AJM, et al. The ScotCap registry: An evaluation of 1000 colon capsule endoscopy procedures carried out in Scotland. Colorectal Disease. John Wiley and Sons Inc; 2025;27. 10.1111/codi.17271

28. Blanes-Vidal V, Baatrup G, Nadimi ES. Addressing priority challenges in the detection and assessment of colorectal polyps from capsule endoscopy and colonoscopy in colorectal cancer screening using machine learning. Acta Oncol (Madr). 2019;58:S29–36. 10.1080/0284186X.2019.1584404

29. Blanes-Vidal V, Nadimi ES, Buijs MM, Baatrup G. Capsule endoscopy vs. colonoscopy vs. histopathology in colorectal cancer screening: matched analyses of polyp size, morphology, and location estimates. Int J Colorectal Dis. 2018;33:1309–12. 10.1007/s00384-018-3064-0

30. Butterly LF, Chase MP, Pohl H, Fiarman GS. Prevalence of Clinically Important Histology in Small Adenomas. Clinical Gastroenterology and Hepatology. 2006;4:343–8. 10.1016/j.cgh.2005.12.021

31. von Renteln D, Pohl H. Polyp Resection -Controversial Practices and Unanswered Questions. Clin Transl Gastroenterol. 2017;8:e76. 10.1038/ctg.2017.6

32. Baatrup G, Bjorsum-Meyer T, Kaalby L, Schelde-Olesen B, Kobaek-Larsen M, Koulaouzidis A, et al. Choice of colon capsule or colonoscopy versus default colonoscopy in FIT positive patients in the Danish screening programme: A parallel group randomised controlled trial. Gut. BMJ Publishing Group; 2025; 10.1136/gutjnl-2024-333687

