## Supplemental Material for "Cost-Effectiveness and Cost-Utility of a Colon Capsule Endoscopy in a Population-Based Screening Program for Colorectal Cancer"

Supplementary Appendix

### SUPPLEMENTARY TABLES

#### Table S1. Background mortality

Yearly incidence for background mortality. Central, eastern and western Europe, 2017-2019 (pre-pandemics)

| | **Mean** | **Min** | **Max** |
|---|---|---|---|
| 50-54 years | 0.567% | 0.288% | 0.853% |
| 55-59 years | 0.864% | 0.462% | 1.199% |
| 60-64 years | 1.304% | 0.737% | 1.753% |
| 65-69 years | 1.892% | 1.121% | 2.496% |
| 70-74 years | 2.702% | 1.728% | 3.473% |
| 75-79 years | 4.609% | 2.933% | 6.016% |
| 80-84 years | 7.477% | 5.178% | 8.742% |
| 85-89 years | 12.788% | 10.065% | 14.226% |
| 90-94 years | 20.756% | 17.929% | 22.383% |
| 95+ years | 30.068% | 27.050% | 32.612% |
| **Total general** | **8.303%** | **0.288%** | **32.612%** |

Source: Global Burden of Disease Collaborative Network. Global Burden of Disease Study 2021 [1].

Table S2. Probabilities associated with the natural history of the disease

| | Case base<br>Average (range) | Source(s) |
|---|---|---|
| **Polyp prevalence** | | |
| Any polyp | 0.27 (0.21 – 0.33) | [2–5] |
| Non-advanced polyp | 0.24 (0.19 – 0.29) | [6] |
| Advanced polyp | 0.032 (0.025 – 0.004) | [6] |
| **CRC prevalence** | | |
| Overall CRC | 0.0059 | [7] |
| Localized CRC | 0.0029 | [8] |
| Regional CRC | 0.0019 | [8] |
| Distant CRC | 0.0011 | [8] |
| **Disease progression** | | |
| Any polypectomy to advanced polyps | 0.176 | [9] |
| Any polypectomy to low-risk polyps | 0.07 | [9] |
| APL to local CRC | 0.034 | [9] |
| Local to regional CRC | 0.6195 | [9] |
| Regional CRC to distant CRC | 0.865 | [9] |
| Healthy to NAPL | 0.012 | [9] |
| No polypoid precursor to CRC | Age specific, 0.003-0.056 | [6] |
| NAPL to APL | 0.024 (0.01-0.04) | [9] |
| Local CRC to Death | 0.029 (0.14-0.044) | [10] |
| Regional CRC to Death | 0.08 | [10] |
| Distal CRC to Death | 0.19 | [10] |

Table S3. Test performance

| Parameter | Stage | Test | Base Case Value (Range) | Source(s) |
|---|---|---|---|---|
| Specificity | Healthy | FIT | 0.964 (0.958-0.969) | [11] |
| | | OC | 1.00 | [11] |
| | | CCE | 0.85 | [12,13] |
| | | CCE-AI | 0.95 | [14] |
| Sensitivity | non-advanced lesions | FIT | 0.076 (0.0607-0.086) | [11] |
| | | OC | 0.85 (0.850-0.950) | [9,15,16] |
| | | CCE | 0.66 – 0.94 | [13,17] |
| | | CCE-AI | 0.87 – 0.96 | [14,17] |
| | advanced lesions | FIT | 0.238 (0.208-0.270) | [11] |
| | | OC | 0.90 (0.900-0.970) | [9,15,16] |
| | | CCE | 93% | [18] |
| | | CCE-AI | 97% | [18] |
| | CRC | FIT | 0.73 (0.603-0.839) | [11] |
| | | OC | 0.95 | [15] |
| | | OC | 0.95 (0.900-0.970) | [16] |
| | | CCE | 86% | [17] |
| | | CCE-AI | 98% | [14] |

Table S4. Complications of optical colonoscopy

| Parameter | Base Case Value (Range)[1] | |
|---|---|---|
| Colonoscopy major hemorrhage rate, % | 0.08 (0.05-0.14) | [19] |
| Colonoscopy perforation rate, % | 0.04 (0.02-0.05) | [19] |
| Mortality rate peer perforation case, % | 7.5 (4.5-16) | [20] |

[1]For clarity, values are presented in percentages instead of proportions.

Table S5. Adherence to tests within the screening program.

| | Base Case Value (Range) | Additional reference |
|---|---|---|
| First FIT of screening campaign | 0.52 (0.26-0.62) | [21,22] |
| Subsequent FIT of screening campaign | 0.47 (0.164 - 0.8)* | [23,24] |
| First OC after positive FIT | 0.80 (0.60-0.90 | [25,26] |
| Follow-up OC after polyp removal | 0.80 (0.60-0.90) | [26] |
| Follow-up OC for a positive screening test | 0.80 (0.50-0.90) | [26] |
| CCE after a positive result in FIT | 0.9 | Assumption |

Based on specific data on this adherence value in Denmark, adherence to subsequent FIT in Denmark was set to 0.6, as reported by the National healthcare Quality Institute in Denmark:

https://www.sundk.dk/media/auioalde/dts-aarsrapport_2024_offentliggjort_version_20251212.pdf

Table S6. Costs according to country

| | Spain | Denmark | Scotland |
|---|---|---|---|
| Invitation | 2.50€ | | |
| GP visit | 73.38€ | | |
| Gastroenterology visit | 242.46€ | | |
| Tests | | | |
| FIT | 4.53€ | 54.00€ | 5.39€ |
| CCE | 520.00€ | 1,684.22€ | 888.93€ |
| Colonoscopy | 316.16€ | 792.18€ | 442.68€ |
| Colonoscopy with biopsy | 354.33€ | 1,350.18€ | 862.75€ |
| Complications | 5,157.00€ | | 4,552.94€ |
| Complications HR | | 1,350.18€ | 717.57€ |
| Complications Perforation | 4,770.59€ | 16,633.43€ | 3,835.37€ |
| Retention | | | |
| Treatments | | | |
| Stage 1 (local) | | | |
| Stage 1 (local) | 10,442.00€ | 8,559.12€ | 12,710.39€ |
| Stage 2 (local) | 11,653.00€ | 8,559.12€ | 12,710.39€ |
| Stage 3 (regional) | 11,688.00€ | 19,553.22€ | 21,433.09€ |
| Stage 4 (distant) | 19,423.00€ | 32458.34361 | 23,075.29€ |
| **Sources** | [27–29] | [30] | [31–33] |

### SUPPLEMENTARY FIGURES

**Figure S1.** Model input for the colonoscopy (A) and CCE (B) pathways

A

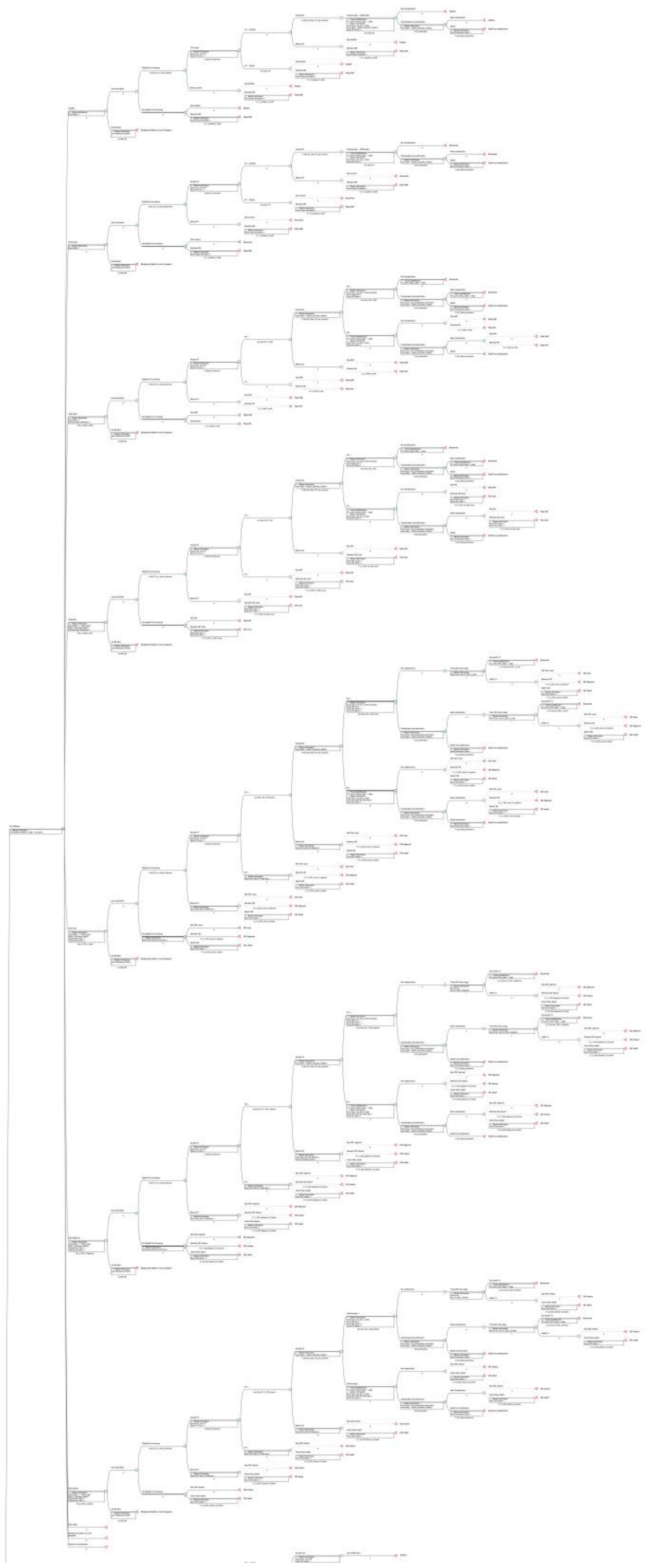

B

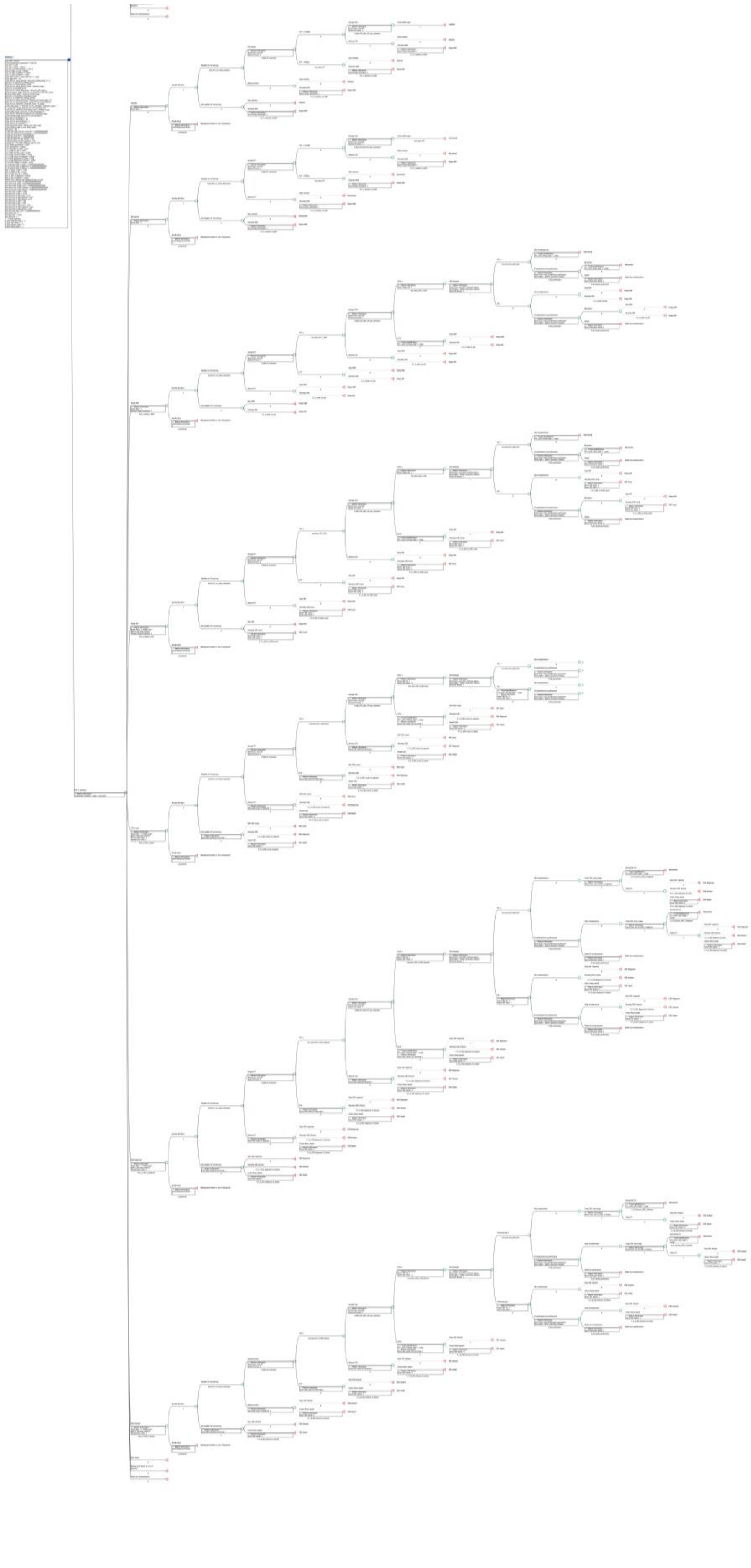

Figure S2. Causes of undetected colorectal cancer in the colon capsule endoscopy (CCE) and optical colonoscopy pathways.

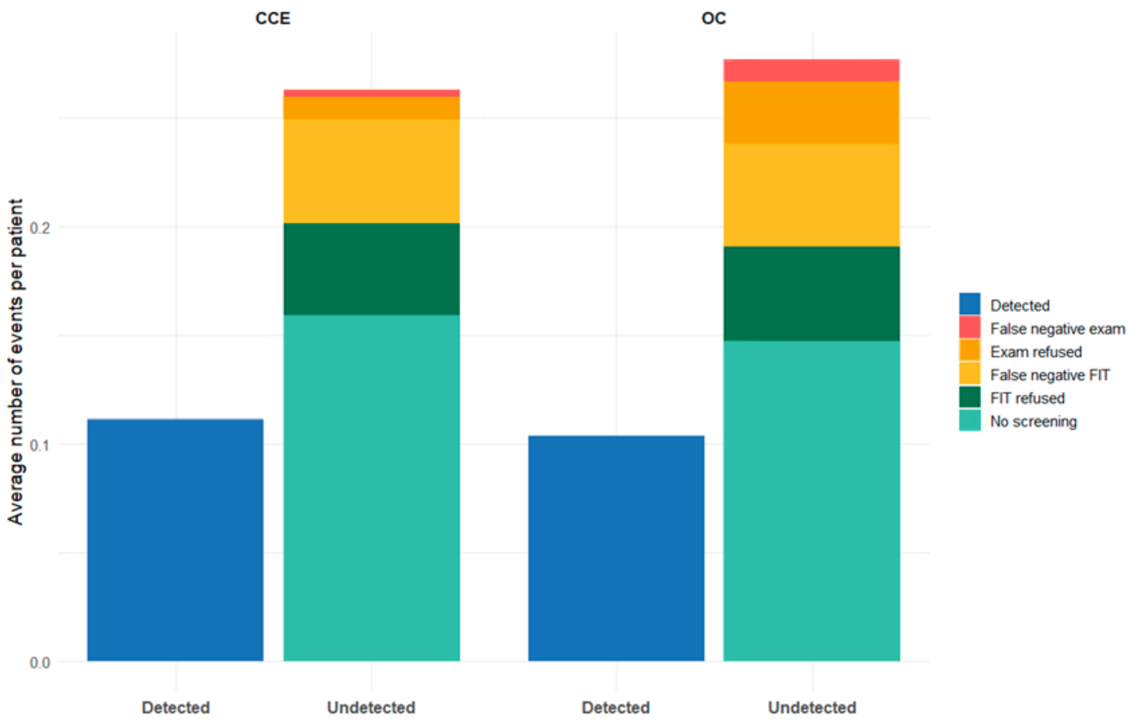

#### REFERENCES

1. Global Burden of Disease Collaborative Network. Global Burden of Disease Study 2021 (GBD 2021) Results [Internet]. Seattle, United States: Institute for Health Metrics and Evaluation (IHME), 2022. 2022 [cited 2025 Apr 16]. https://vizhub.healthdata.org/gbd-results/. Accessed 16 Apr 2025

2. Vatn MH, Stalsberg H. The prevalence of polyps of the large intestine in Oslo: An autopsy study. Cancer. 1982;49:819–25. https://doi.org/10.1002/1097-0142(19820215)49:4<819::AID-CNCR2820490435>3.0.CO;2-D

3. Paspatis GA, Papanikolaou N, Zois E, Michalodimitrakis E. Prevalence of polyps and diverticulosis of the large bowel in the Cretan population. An autopsy study. Int J Colorectal Dis. 2001;16:257–61. https://doi.org/10.1007/s003840100304

4. Johannsen LG, Momsen O, Jacobsen NO. Polyps of the large intestine in Aarhus, Denmark. An autopsy study. Scand J Gastroenterol [Internet]. 1989;24:799–806. https://doi.org/10.3109/00365528909089217

5. Williams AR, Balasooriya BA, Day DW. Polyps and cancer of the large bowel: a necropsy study in Liverpool. Gut [Internet]. 1982;23:835–42. https://doi.org/10.1136/gut.23.10.835

6. Ladabaum U, Song K. Projected national impact of colorectal cancer screening on clinical and economic outcomes and health services demand. Gastroenterology. W.B. Saunders; 2005;129:1151–62. https://doi.org/10.1053/j.gastro.2005.07.059

7. Ferlay J, Ervik M, Lam F, Colombet M, Mery L, Piñeros M. Global Cancer Observatory: Cancer Today. Lyon: International Agency for Research on Cancer [Internet]. 2023 [cited 2025 Apr 16]. https://gco.iarc.fr/today/en/dataviz/tables-prevalence?mode=cancer&types=2&cancers=41&populations=908&multiple_populations=1&age_start=10&group_populations=1. Accessed 16 Apr 2025

8. Dyba T, Randi G, Bray F, Martos C, Giusti F, Nicholson N, et al. The European cancer burden in 2020: Incidence and mortality estimates for 40 countries and 25 major cancers. Eur J Cancer. Elsevier Ltd; 2021;157:308–47. https://doi.org/10.1016/j.ejca.2021.07.039

9. Lopes L, Certo M, Veiga P, Canena. J. Cost-effectiveness of Colorectal Screening in a European Country. A Comparison of Five Alternative Screening Strategies. J Surg Res (Houst). Fortune Journals; 2022;05. https://doi.org/10.26502/jsr.10020253

10. Digestive Cancers Europe. White paper: Colorectal Screening in Europe: Saving Lives & Saving Money [Internet]. 2019. https://digestivecancers.eu/publication/colorectal-screening-in-europe/. Accessed 31 Jan 2025

11. Imperiale TF, Ransohoff DF, Itzkowitz SH, Levin TR, Lavin P, Lidgard GP, et al. Multitarget Stool DNA Testing for Colorectal-Cancer Screening. New England Journal of Medicine. New England Journal of Medicine (NEJM/MMS); 2014;370:1287–97. https://doi.org/10.1056/nejmoa1311194

12. Rokkas T, Papaxoinis K, Triantafyllou K, Ladas SD. A meta-analysis evaluating the accuracy of colon capsule endoscopy in detecting colon polyps. Gastrointest Endosc. 2010;71:792–8. https://doi.org/10.1016/j.gie.2009.10.050

13. Vuik FER, Nieuwenburg SAV, Moen S, Spada C, Senore C, Hassan C, et al. Colon capsule endoscopy in colorectal cancer screening: A systematic review. Endoscopy. Georg Thieme Verlag; 2021;53:815–24. https://doi.org/10.1055/a-1308-1297

14. Moen S, Vuik FER, Kuipers EJ, Spaander MCW. Artificial Intelligence in Colon Capsule Endoscopy—A Systematic Review. Diagnostics. Multidisciplinary Digital Publishing Institute (MDPI); 2022. https://doi.org/10.3390/diagnostics12081994

15. Gupta S, Jacobs ET, Baron JA, Lieberman DA, Murphy G, Ladabaum U, et al. Risk stratification of individuals with low-risk colorectal adenomas using clinical characteristics: A pooled analysis. Gut. BMJ Publishing Group; 2017;66:446–53. https://doi.org/10.1136/gutjnl-2015-310196

16. Ladabaum U, Mannalithara A, Meester RGS, Gupta S, Schoen RE. Cost-Effectiveness and National Effects of Initiating Colorectal Cancer Screening for Average-Risk Persons at Age 45 Years Instead of 50 Years. Gastroenterology. W.B. Saunders; 2019;157:137–48. https://doi.org/10.1053/j.gastro.2019.03.023

17. Sulbaran M, Bustamante-Lopez L, Bernardo W, Sakai CM, Sakai P, Nahas S, et al. Systematic review and meta-analysis of colon capsule endoscopy accuracy for colorectal cancer screening. An alternative during the Covid-19 pandemic? J. Med. Screen. SAGE Publications Ltd; 2022. p. 148–55. https://doi.org/10.1177/09691413221074803

18. Alihosseini S, Aryankhesal A, Sabermahani A. Second-generation colon capsule endoscopy for detection of colorectal polyps: A meta-analysis. Med J Islam Repub Iran. Iran University of Medical Sciences; 2020;34:1–8. https://doi.org/10.34171/mjiri.34.81

19. Baile-Maxía S, Mangas-Sanjuán C, Ladabaum U, Hassan C, Rutter MD, Bretthauer M, et al. Risk Factors for Metachronous Colorectal Cancer or Advanced Adenomas After Endoscopic Resection of High-risk Adenomas. Clinical Gastroenterology and Hepatology. W.B. Saunders; 2023. p. 630–43. https://doi.org/10.1016/j.cgh.2022.12.005

20. Click B, Pinsky PF, Hickey T, Doroudi M, Schoen RE. Association of colonoscopy adenoma findings with long-term colorectal cancer incidence. JAMA - Journal of the American Medical Association. American Medical Association; 2018;319:2021–31. https://doi.org/10.1001/jama.2018.5809

21. Hoffman RM, Steel S, Yee EFT, Massie L, Schrader RM, Murata GH. Colorectal cancer screening adherence is higher with fecal immunochemical tests than guaiac-based fecal occult blood tests: A randomized, controlled trial☆. Prev Med (Baltim). 2010;50:297–9. https://doi.org/10.1016/j.ypmed.2010.03.010

22. Hassan C, Rossi PG, Camilloni L, Rex DK, Jimenez-Cendales B, Ferroni E, et al. Meta-analysis: adherence to colorectal cancer screening and the detection rate for advanced neoplasia, according to the type of screening test. Aliment Pharmacol Ther. 2012;36:929–40. https://doi.org/10.1111/apt.12071

23. Murphy CC, Sen A, Watson B, Gupta S, Mayo H, Singal AG. A Systematic Review of Repeat Fecal Occult Blood Tests for Colorectal Cancer Screening. Cancer Epidemiology, Biomarkers & Prevention. 2020;29:278–87. https://doi.org/10.1158/1055-9965.EPI-19-0775

24. Castells A, Quintero E, Bujanda L, Castán-Cameo S, Cubiella J, Díaz-Tasende J, et al. Effect of invitation to colonoscopy versus faecal immunochemical test screening on colorectal cancer

mortality (COLONPREV): a pragmatic, randomised, controlled, non-inferiority trial. The Lancet. 2025;405:1231–9. https://doi.org/10.1016/S0140-6736(25)00145-X

25. Wong CK, Lam CL, Wan Y, Fong DY. Cost-effectiveness simulation and analysis of colorectal cancer screening in Hong Kong Chinese population: comparison amongst colonoscopy, guaiac and immunologic fecal occult blood testing. BMC Cancer. 2015;15:705. https://doi.org/10.1186/s12885-015-1730-y

26. Frazier AL, Colditz GA, Fuchs CS, Kuntz KM. Cost-effectiveness of screening for colorectal cancer in the general population. JAMA [Internet]. 2000;284:1954–61. https://doi.org/10.1001/jama.284.15.1954

27. Valcárcel-Nazco C G-PLR-SAH-YAT-CAG-PHA-SAH-REG-FCG-RDG-CEG-SMA. Análisis de la efectividad clínica y coste-efectividad de la ampliación a 74 años del cribado del cáncer colorrectal en la población general. Informes de Evaluación de Tecnologías Sanitarias. [Internet]. 2023. https://www3.gobiernodecanarias.org/sanidad/scs/content/b29017e1-346a-11ef-ae6a-69e2085c71b4/SESCS_2023_CRIBADO_CCR_1.pdf. Accessed 6 Feb 2025

28. Suárez Ferrer C, González Lama Y, Blázquez Gómez I, Barrios Peinado C, Martinez Porras JL, Vera Mendoza MI, et al. Utilidad y coste de la cápsula endoscópica. Tres años de experiencia de nuestro centro. Gastroenterol Hepatol. 2013;36:121–6. https://doi.org/10.1016/j.gastrohep.2012.10.009

29. Ibarrondo O, Lizeaga G, Martínez-Llorente JM, Larrañaga I, Soto-Gordoa M, Álvarez-López I. Health care costs of breast, prostate, colorectal and lung cancer care by clinical stage and cost component. Gac Sanit [Internet]. Ediciones Doyma, S.L.; 2022;36:246–52. https://doi.org/10.1016/j.gaceta.2020.12.035

30. Skovlund LK. Cost-Effectiveness Analysis of Colon Capsule Endoscopy in Colorectal Cancer Screening. Aalborg University; 2020.

31. Westwood M, Ramos IC, Lang S, Luyendijk M, Zaim R, Stirk L, et al. Faecal immunochemical tests to triage patients with lower abdominal symptoms for suspected colorectal cancer referrals in primary care: A systematic review and cost-effectiveness analysis. Health Technol. Assess. (Rockv). NIHR Journals Library; 2017. https://doi.org/10.3310/hta21330

32. Picot J, Rose M, Cooper K, Pickett K, Lord J, Harris P, et al. Virtual chromoendoscopy for the real-time assessment of colorectal polyps in vivo: A systematic review and economic evaluation. Health Technol Assess (Rockv). NIHR Journals Library; 2017;21:1–307. https://doi.org/10.3310/hta21790

33. Innovative Medical Technology Overview. Colon capsule endoscopy (CCE) for the detection of colorectal polyps and cancer [Internet]. [Internet]. 2024. https://shtg.scot/media/2430/20240109-cce-update-v10.pdf. Accessed 3 Apr 2025
